# Inter-shot Motion Correction of Segmented 3D-GRASE ASL Perfusion Imaging with Self-Navigation and CAIPI

**DOI:** 10.1101/2025.09.18.25335960

**Authors:** Minhao Hu, Frederik J. Lange, Peter Jezzard, Joseph G. Woods, Mark Chiew, Thomas W. Okell

## Abstract

**Purpose:** Segmented 3D Gradient and Spin Echo (GRASE) is commonly used in Arterial Spin Labeling (ASL) perfusion imaging. However, it is vulnerable to inter-shot motion, leading to subtraction errors that cannot be corrected. We developed a retrospective self-navigated inter-shot motion correction method for segmented 3D-GRASE ASL imaging with Controlled Aliasing in Parallel Imaging (CAIPI).

**Methods:** Multiple shots, each uniformly covering k-space at distinct sample locations, allow a self-navigator image to be reconstructed using SENSE for each shot. Rigid-body motion estimation across the self-navigators is incorporated into a motion-compensated forward model for image reconstruction. To support self-navigation, two CAIPI-sampled segmented 3D-GRASE trajectories that ensure full k-space coverage were explored for point spread function (PSF) profiles and g-factor effects. Our approach was evaluated against conventional inter-volume registration and a previously proposed method, alignedSENSE. Additionally, we compared tag-control interleaving strategies to assess their impact on motion robustness in five healthy volunteers with instructed head motion.

**Results:** Our method effectively reduced motion artifacts and outperformed conventional inter-volume correction by 12.3% in correlation coefficient, 4.5% in Structural Similarity Index Measure (SSIM), and 40.1% in temporal SNR. It matched alignedSENSE performance while requiring only 20% of the computational time. All evaluated CAIPI sampling variants enabled robust motion correction, although tradeoffs were observed between through-plane blurring and SNR performance. The tag-control (T/C) inner loop acquisition yielded better motion robustness across all quantitative metrics.

**Conclusion:** Self-navigated inter-shot motion correction using CAIPI sampling and a T/C inner loop for segmented 3D-GRASE ASL can improve image quality and motion robustness.

## 1. Introduction

Arterial Spin Labeling (ASL) is a non-invasive MRI technique for quantifying cerebral blood flow (CBF), which uses magnetically labeled arterial blood water as an endogenous tracer^1^. By eliminating the need for exogenous contrast agents, ASL is particularly suitable for populations with contraindications to contrast-based agents, such as patients with renal failure. ASL provides quantitative estimates of CBF, allowing more robust comparisons across brain regions, subjects and in longitudinal studies^2^. Therefore, ASL has been adopted in both clinical and research settings for evaluating cerebral perfusion and detecting perfusion abnormalities associated with various neurological conditions^3^.

However, the perfusion signal derived from the subtraction of tag and control images differs by only about 1–2% of the signal intensity of static tissue^4^. Therefore, SNR is inherently low for ASL and multiple repetitions are required for averaging. It is also highly sensitive to motion, as even small misalignments, including between individual shots within a single volume, can introduce large subtraction errors and result in inaccurate CBF estimation^5^. Single-shot 3D acquisitions can mitigate motion artifacts, but the spatial coverage will be limited, or the echo train will be prolonged, leading to T2-blurring in the through-plane direction^4,6^. To address this issue, segmented (multi-shot) 3D acquisitions e.g. segmented 3D-GRASE^7^ or 3D RARE stack of spirals^8^ are recommended in ASL consensus papers^4,9^. Segmented 3D acquisitions allow shorter echo trains by dividing k-space into multiple segments, thereby mitigating T2-blurring while maintaining high SNR efficiency, whole-brain coverage and compatibility with background suppression. However, segmentation also leads to a multiplication of the total acquisition time and introduces the risk of inter-shot motion, i.e. subject motion occurring between successive segments. Unlike inter-volume motion artifacts, such *intra-volume* inter-shot motion artifacts cannot be corrected using standard image registration methods in conventional ASL post-processing pipelines^10^. Developing effective inter-shot motion correction methods is therefore critical for realizing the full potential of segmented 3D ASL acquisitions.

Prospective motion correction methods aim to reduce inter-volume and inter-shot motion artifacts by incorporating low-resolution navigators^11,12^ or external optical tracking systems^13^ to adjust imaging parameters or reacquire^14^ corrupted data in real time during the scan. While effective, these approaches often require substantial modifications to the pulse sequence or additional hardware and may lead to increased total acquisition time. As a more practical alternative, Tan et al.^15^ incorporated 3D-GRASE with a widely used retrospective motion correction method PROPELLER^16^. Huber et al.^17^ further improved this by jointly estimating motion and geometric distortion. However, these methods suffered from suboptimal SNR efficiency and were limited to 2D motion detection. Highton et al.^18^ proposed an interleaved segmented 3D-GRASE readout where each shot comprised SENSE-style undersampled k-space data. They used SENSE for individual shot reconstruction and corrected inter-shot motion via rigid image registration. However, by applying registration on images rather than incorporating motion parameters into the reconstruction, their approach introduced interpolation artifacts and higher g-factor noise amplification penalties. Spann et al.^6^ proposed a robust single-shot 3D-GRASE readout with time-dependent 2D CAIPIRINHA sampling and incorporated spatio-temporal total generalized variation regularization to suppress motion artifacts. While this offered improved robustness compared to standard segmented 3D-GRASE, it did not explicitly correct inter-shot rigid motion.

AlignedSENSE^19^ is a widely used retrospective motion correction method that has been applied across various imaging modalities and readout schemes^20^. It incorporates motion directly into the forward model and jointly estimates the motion-free image and motion parameters using an alternating optimization approach.

Inspired by Spann et al. and alignedSENSE, our study proposes a self-navigated inter-shot motion correction method for segmented 3D-GRASE ASL with Controlled Aliasing in Parallel Imaging (CAIPI) sampling. **Figure 1** illustrates the overview of the proposed method. Unlike the approach by Highton et al.^18^, the segmented 3D-GRASE readout employs interleaved CAIPI sampling patterns, with each segment acquired in a single shot. This design reduces the echo train length, thereby mitigating T2 blurring effects. After acquiring all segments, the data can be combined to form a fully sampled k-space volume, which can be used for sensitivity map estimation. Meanwhile, each individual shot uniformly covers the entire k-space and so can be reconstructed using SENSE to produce low-SNR self-navigator images, which are nonetheless sufficient for motion estimation via image registration. The estimated inter-shot rigid motion is then incorporated into a final motion-compensated reconstruction, enabling recovery of a high-quality motion-free image from the multi-shot data without noise amplification or interpolation penalties.

**Figure 1.**
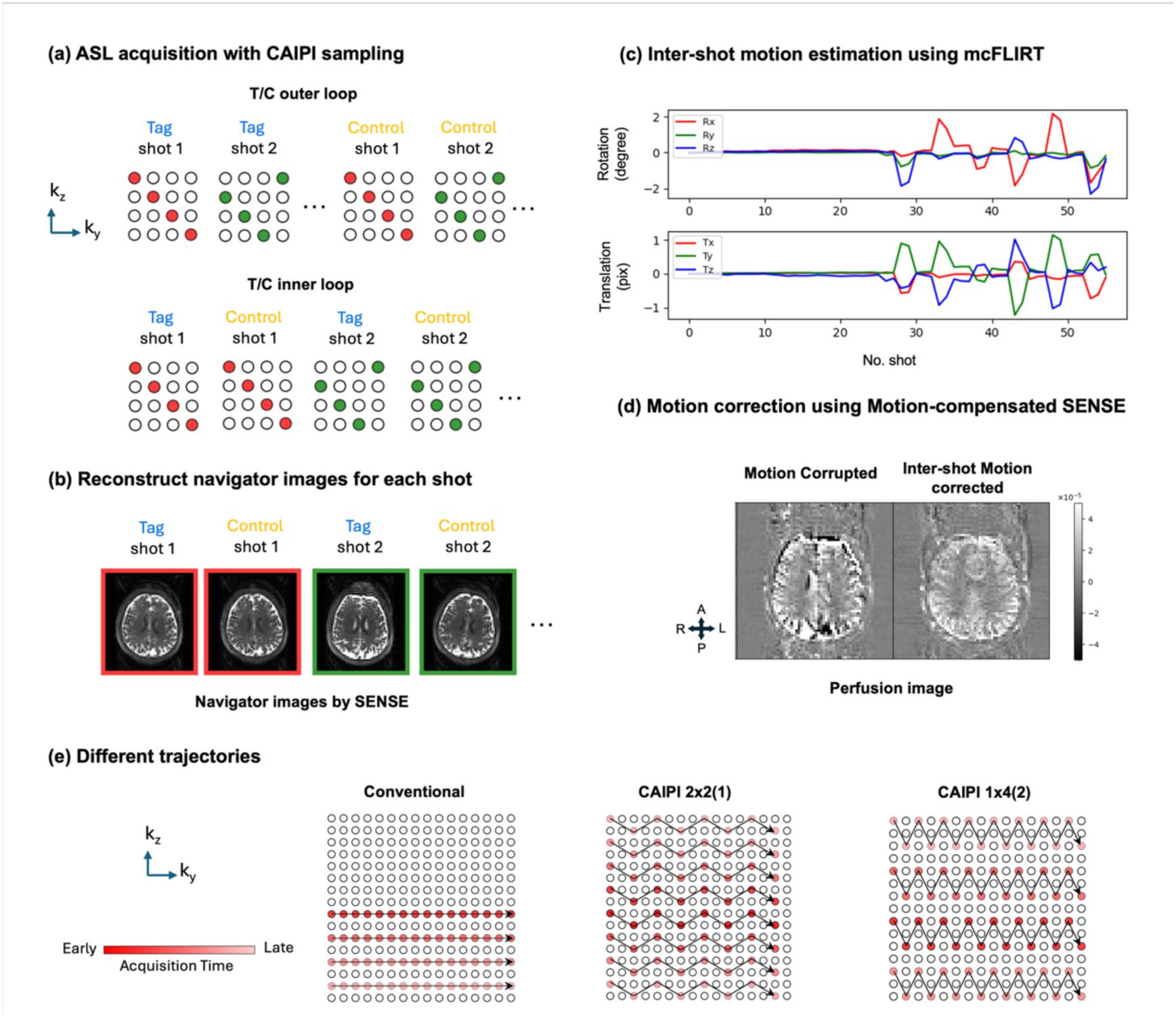
**Overview of the proposed approach**. (a) The diagram illustrates how segmented k-space data are acquired across shots over time, with different colors indicating k-space lines sampled in different shots and under different tag/control conditions. The T/C inner loop alternates tag and control modes before switching sampling patterns, while the T/C outer loop does so after acquiring each volume. (b) Navigator images reconstruction using conventional SENSE per shot for both tag and control conditions. (c) Translation and rotation estimation across shots and tag/control conditions obtained via mcFLIRT. (d) Reconstruction of motion-corrected images using a motion-compensated model. (e) The schematic shows different trajectories including a conventional approach and two CAIPI variants. Red spots represent the acquired k-space lines (illustrated for the first shot only), with transparency reflecting acquisition timing (darker means earlier). One solid arrow indicates the trajectory within a single spin echo. For clarity, only a 16 x 16 portion in the phase-encoding (y) and partition (z) directions is shown, whereas the actual matrix size is 64 x 32.

In this study we demonstrated using in vivo data from five healthy volunteers that this method outperformed conventional segmented 3D-GRASE in the presence of inter-shot motion. Two CAIPI sampling strategies were evaluated against a conventional trajectory, and three motion correction approaches were compared both quantitatively and qualitatively. Additionally, we investigated the impact of two different tag-control interleaving schemes to establish whether placing the tag-control interleaving in the innermost loop was more motion-robust in practice.

## 2. Methods

### 2.1 Trajectory design

Some conventional implementations acquire only partial k-space per shot, such as center-out half-partition sampling. While such designs preserve the point spread function (PSF) and reduce T2-related blurring, the incomplete k-space coverage per shot limits the feasibility of reconstructing reliable self-navigator images. Alternatively, other implementations used SENSE-style undersampling patterns which provided the potential for navigator reconstruction but suffered from higher g-factor penalties and reduced SNR efficiency^18^.

To address this limitation, we propose integrating CAIPI-style sampling into the segmented 3D-GRASE acquisition. This produces uniform undersampling patterns characterized by 𝑅𝑦 × 𝑅𝑧 (𝛥𝑧), where 𝑅𝑦 and 𝑅𝑧 are acceleration factors in the in-plane and through-plane phase encoding directions, respectively, and 𝛥𝑧 is the partition shift applied between successive k-space lines^21^. By shifting the CAIPI pattern across shots, full k-space coverage can be achieved after 𝑅 = 𝑅𝑦 × 𝑅𝑧 repetitions, allowing both high-quality final reconstruction and low-SNR navigator reconstruction from individual shots.

To ensure compatibility with the 3D-GRASE readout, which comprised multiple spin echoes (defined by the turbo factor) and multiple EPI gradient echoes between each spin echo (defined by the EPI factor), the CAIPI sampling was designed such that:

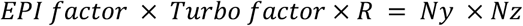

Where 𝑁𝑦 and 𝑁𝑧 were the number of lines acquired in the phase encoding and partition encoding directions, respectively. In addition to satisfying this constraint, we optimized the sampling to minimize PSF sidelobes and reduced sensitivity to off-resonance artifacts by ensuring smooth variation of signal intensity and phase accrual across both phase-encoding and partition directions.

In this study, we set the matrix size to 𝑁𝑦 = 64 and 𝑁𝑧 = 32, with an effective acceleration factor for each shot of 𝑅 = 4. As a reference, we adopted a conventional segmented 3D-GRASE trajectory, in which we acquired one full partition per spin echo and one phase-encoding line per EPI gradient echo, i.e. an EPI factor of 𝑁𝑦 = 64 and a Turbo factor of 𝑁𝑧/𝑅 = 8. In this configuration, each shot sampled every other partition in half of 3D k-space. Based on these parameters and constraints described above, we designed and evaluated two CAIPI sampling variants. Detailed acquisition parameters for each strategy are listed in **Table 1**.

**Table 1.** Imaging parameters for the different trajectories. Turbo factor denotes the number of spin echoes per shot, while EPI factor is the number of phase encoding lines within each spin echo. TR indicates the interval between two ASL preparation pulses.

|  | Conventional | CAIPI 2x2(1) | CAIPI 1x4(2) |
| --- | --- | --- | --- |
| Turbo Factor | 8 | 16 | 8 |
| EPI Factor | 64 | 32 | 64 |
| TE (ms) | 37.86 | 22.52 | 39.16 |
| TR (ms) | 3990 | 4030 | 4000 |
| Echo Spacing (ms) | 0.50 | 0.52 | 0.54 |

𝐶𝐴𝐼𝑃𝐼 1 × 4 (2): Maintained the same EPI and Turbo factors as the conventional trajectory, with four partitions sampled per spin echo. This design preserved the original echo train length while introducing controlled aliasing along the partition direction.

𝐶𝐴𝐼𝑃𝐼 2 × 2 (1): Reduced the EPI factor by half while doubling the Turbo factor to maintain full k-space coverage. While this resulted in a slightly longer readout time, it potentially improves PSF uniformity and may offer greater robustness to off-resonance effects due to a larger step size (Δ𝑘) in the phase-encoding direction.

In addition to sampling density, the order of k-space traversal played a critical role in determining the PSF and sensitivity to motion and off-resonance effects. All proposed trajectories employed symmetric center-out partition ordering to minimize T2-weighting and maximized SNR by acquiring central k-space early in the echo train. Phase-encoding and partition-encoding blipped between refocusing pulses were carefully constrained to ensure smooth phase evolution and T2 decay weighting, reducing artefacts that would result from abrupt changes in signal intensity or phase.

PSF simulations were conducted under on-and off-resonance (𝛥𝑓 = 100 𝐻𝑧) conditions to compare the proposed CAIPI trajectories with the conventional segmented acquisition. The simulations were performed using T2=110 ms, T2*=66 ms and a refocusing flip angle of 180° for simplicity.

g-factor maps were estimated using the pseudo-multiple replica method^22^. Multiple realizations of Gaussian noise were added to the k-space data, followed by SENSE reconstruction of both fully sampled and undersampled (R = 4) data. Voxelwise g-factor values were then computed as the ratio of standard deviations across noise realizations between the two datasets, normalized by square root of the ^acceleration^ factor R.

### 2.2 Inter-shot Motion Correction

The proposed motion correction pipeline consisted of three main stages: navigator image reconstruction, inter-shot motion estimation, and motion-compensated image reconstruction. This framework took advantage of the full k-space coverage enabled by the proposed CAIPI sampling scheme, allowing per-shot navigator image reconstruction for accurate motion estimation. A total of 𝑁 pairs of tag and control volumes were acquired, with each volume segmented into 𝑅 shots so that each shot sampled a unique CAIPI pattern. This ensured that R shots can be combined to form a fully sampled k-space volume, which was essential for both sensitivity map estimation and motion-compensated reconstruction. In addition, the use of CAIPI sampling reduced noise amplification by minimizing g-factor penalties, thereby improving the SNR of both the navigator images and the final motion-corrected reconstruction.

#### 2.2.1 Navigator Image Reconstruction

Utilizing the full k-space coverage per shot, each individual shot was reconstructed to generate a self-navigator image for motion estimation. Unlike joint estimation approaches such as alignedSENSE, this strategy leveraged the well-conditioned nature of the CAIPI undersampled data to achieve reliable motion estimates through conventional image registration, while avoiding the computational complexity and parameter tuning challenges associated with iterative joint optimization methods.

For each shot, an undersampled image was reconstructed using SENSE with l2-regularization.

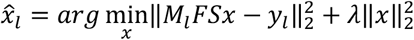

Where 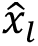 denotes the reconstructed self-navigator image for the 𝑙-th shot (𝑙 ∈ [1, 𝑅]), 𝑦_!_is the corresponding acquired k-space data, 𝑀_l_ is the undersampling mask for 𝑙-th shot, 𝐹 is the Fourier transform operator and 𝑆 is the operator for coil sensitivity maps. 𝜆 is the regularization parameter which is empirically set to 1 × 10^-3^. The optimization problem is solved using the Conjugate Gradient algorithm.

To improve robustness against motion and ensure full k-space coverage in coil sensitivity estimation, all motion-corrupted volumes were temporally averaged before ESPIRiT^23^ calibration. Although subject motion introduces inconsistency across shots, ESPIRiT has been shown to exhibit inherent robustness to moderate motion artifacts due to its calibration region design^18^. In addition, we set the crop threshold to zero, ensuring that the estimated sensitivity maps extended across the entire field of view, including peripheral regions. This strategy helped mitigate the risk of coil sensitivity map truncation caused by large displacements, which could otherwise degrade single-shot navigator reconstruction and downstream motion registration accuracy.

#### 2.2.2 Inter-Shot Motion Estimation

Although minor non-rigid components may exist, head motion is typically modeled as a rigid-body transformation which is widely adopted in previous work^24^. The rigid transform has six degrees of freedom (DOF): three translations (𝑡_x_, 𝑡_y_, 𝑡_z_) and three rotations about the three axes (𝑟_x_, 𝑟_y_, 𝑟_z_).

We denote the rigid-body transformation operator for the 𝑙-th shot as 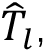 defined by 6-DOF parameter set (𝑡_x_, 𝑡_y_, 𝑡_z_, 𝑟_x_, 𝑟_y_, 𝑟_z_)_l_. These parameters were estimated by registering each navigator image 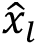 to a reference image 𝑥*_ref_*, which was chosen as the first shot of the first volume, i.e., 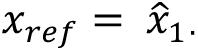. Image registration was performed using mcFLIRT^2324^, with the cost function set to normalized correlation (normcorr) to improve robustness under low-SNR conditions.

#### 2.2.3 Motion-Compensated Reconstruction

The estimated motion parameters are incorporated into a motion-compensated forward model to reconstruct a motion-corrected image by solving the following optimization problem:

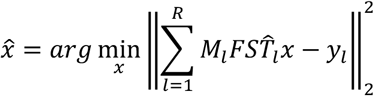

Inspired by alignedSENSE^19^, which also avoids explicit regularization, we leveraged the well-conditioned nature of the motion-compensated encoding operator, due to parallel coil redundancy and full k-space coverage, to achieve a stable and accurate reconstruction using a Conjugate Gradient (CG) optimization. The CG solver was run with a maximum of 200 iterations and convergence was typically reached earlier using a relative residual error tolerance of 1 × 10^-10^.

In addition to inter-shot motion, inter-volume motion can also degrade the final ASL perfusion images. In this study, both inter-shot and inter-volume motions were accounted for by aligning the navigator images from all shots and volumes to a single reference. The resulting motion parameters were integrated into the motion-compensated reconstruction, eliminating the need for extra image registration in post-processing and avoiding interpolation-induced blurring.

### 2.3 Tag-Control Interleaving

As recommended by the ASL consensus papers^4,9^, the tag-control (T/C) loop should be placed in the innermost loop position when using a segmented readout to ensure accurate subtraction. However, this strategy has not been consistently adopted in all studies in the literature and, to the best of our knowledge, its impact on motion robustness has not been thoroughly evaluated.

In this study, we investigated the effect of T/C interleaving loop order on ASL image quality by comparing two configurations (**Figure 1a**): 1) T/C inner loop, where a shot with the same CAIPI sampling pattern was acquired for tag and then control conditions before moving on to the next shot; and 2) T/C outer loop, where all tag shots for a volume were acquired before acquiring the control shots.

The T/C inner loop configuration was hypothesized to enhance motion robustness by minimizing the temporal gap between corresponding tag and control shots, thereby reducing misalignment due to motion, particularly under moderate motion conditions.

### 2.4 Data Acquisition

We modified a previously described 3D-GRASE pseudo-Continuous ASL (pCASL) sequence^25^ on a 3T Prisma scanner (Siemens Healthineers, Erlangen, Germany) equipped with a 32-channel receive-only head coil. The sequence included Water suppression Enhanced through T1 effects (WET)^26^ presaturation and two global background suppression inversion pulses with timing optimized to perfectly null tissues with T1 equal to 700𝑚𝑠 or 1400 ms at a time point 100ms prior to readout excitation.

The pCASL parameters were: tag RF flip angle= 20°, duration= 500𝜇𝑠, mean tag gradient = 0.80𝑚𝑇/𝑚, tag gradient amplitude = 6.0𝑚𝑇/𝑚, labeling duration (LD) = 1800𝑚𝑠, post-label delay (PLD) = 1800𝑚𝑠, 𝑇𝑅_012_ = 4000𝑚𝑠 (time between ASL preparations). The labeling plane was positioned through the proximal V3 segment of the vertebral arteries as per previous studies^27^.

3D-GRASE readout parameters were: refocusing flip angle= 120°, FOV= 230 × 230 × 115 𝑚𝑚³, matrix size= 64 × 64 × 32, voxel size= 3.6 𝑚𝑚 isotropic, bandwidth= 2298 𝐻𝑧/𝑝𝑖𝑥𝑒𝑙, and no slice oversampling. Echo shifting was applied for all trajectories, where applicable, to smooth the signal evolution across shots. Additional readout parameters are summarized in **Table 1**.

Five healthy volunteers (1 female, age 26 ± 1.7 years) were scanned under a technical development protocol approved by local ethics and institutional committees. Subjects remained stationary during the acquisition of 6 motion-free 3D-GRASE volumes (3 tag-control pairs, time 1min 36s) followed by moderate cued head motion during the acquisition of 8 additional volumes (4 tag-control pairs, time 2min8s). The motion was guided by a visual stimulus^28^ to ensure consistency across scans. The total acquisition time for 7 tag-control pairs was 3min 44s. In each session, a T1-weighted anatomical image (1.7 𝑚𝑚 isotropic) was acquired for registration and tissue segmentation (FOV= 220 × 200 × 218, 𝑇𝑅/𝑇𝐸/𝑇𝐼 = 1900/3.71/904𝑚𝑠, flip angle= 8°, acquisition time= 3𝑚 40𝑠).

Acquisitions with three different k-space trajectories were tested in each subject, including one conventional 3D-GRASE reference and two CAIPI sampling variants. All experiments were repeated with both tag-control interleaving strategies: T/C inner loop and T/C outer loop.

### 2.5 Evaluation

A brain mask and a grey matter mask were generated from the T1-weighted image using FSL tools^29^ and then registered to the motion-free ASL reference volume. Perfusion-weighted images 𝐼_3_ were obtained by subtraction between the 𝑡-th pair of tag and control images. To evaluate temporal signal stability, we computed the temporal SNR (tSNR) within a grey matter mask. tSNR is defined as:

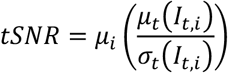

Where 𝑖 indexes voxels within the grey matter mask, 𝜇_3_(·) and 𝜎_3_(·) represent the temporal mean and standard deviation across time for each voxel, and 𝜇_4_(·) denotes the spatial mean across all voxels within the grey matter mask.

To evaluate similarity to a (motion-free) reference image, the Pearson correlation coefficient (Corr) and structural similarity index (SSIM) were calculated between the time-averaged test images 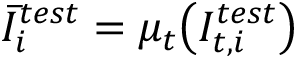 and reference image 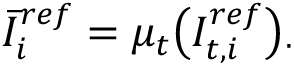 The images were first masked using a brain mask and flattened into vectors for voxel-wise comparison. The Pearson correlation is defined as:

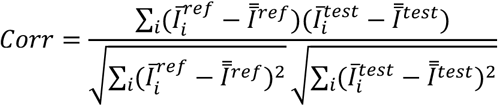

Where 𝑖 indexes voxels within the brain mask and 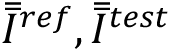 are the spatial mean across voxels, i.e.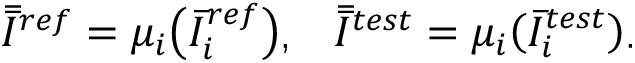

SSIM was computed using a Gaussian kernel with sigma=1.5 and kernel size of 11 voxels isotropic, with constants k1=0.01 and k2=0.03. The SSIM was computed over the brain region on temporal mean images and then averaged across all voxels within the brain mask to obtain a single SSIM score.

When comparing images not already intrinsically registered to the same space, the images were linearly registered with 6 degrees of freedom using FLIRT^24^ and a halfway transformation was applied to match the blurring introduced by interpolation during image registration.

To assess the statistical significance of any improvements achieved by motion correction, paired t-tests were conducted across subjects for all metrics when comparing different methods. A p-value lower than 0.05 was considered statistically significant.

## 3. Results

### 3.1 Trajectory Simulations

To evaluate the effective resolution and noise amplification of the proposed k-space sampling strategies, we simulated the point spread function (PSF) and computed g-factor maps for comparison. **Figure 2** illustrates the PSF profiles along the phase-encoding direction (y-axis) and partition direction (z-axis) under both on-resonance (**Figure 2a**) and 100𝐻𝑧 off-resonance (**Figure 2b**) conditions. Effective resolutions ( 𝐸𝑓𝑓𝑅𝑒𝑠_’_ and 𝐸𝑓𝑓𝑅𝑒𝑠_&_) were quantified as the full width at half maximum (FWHM) of the PSF in each direction. For reference, the ideal effective resolution under 𝑇2 = ∞ conditions would be 100% (normalized to the nominal voxel size).

**Figure 2.**
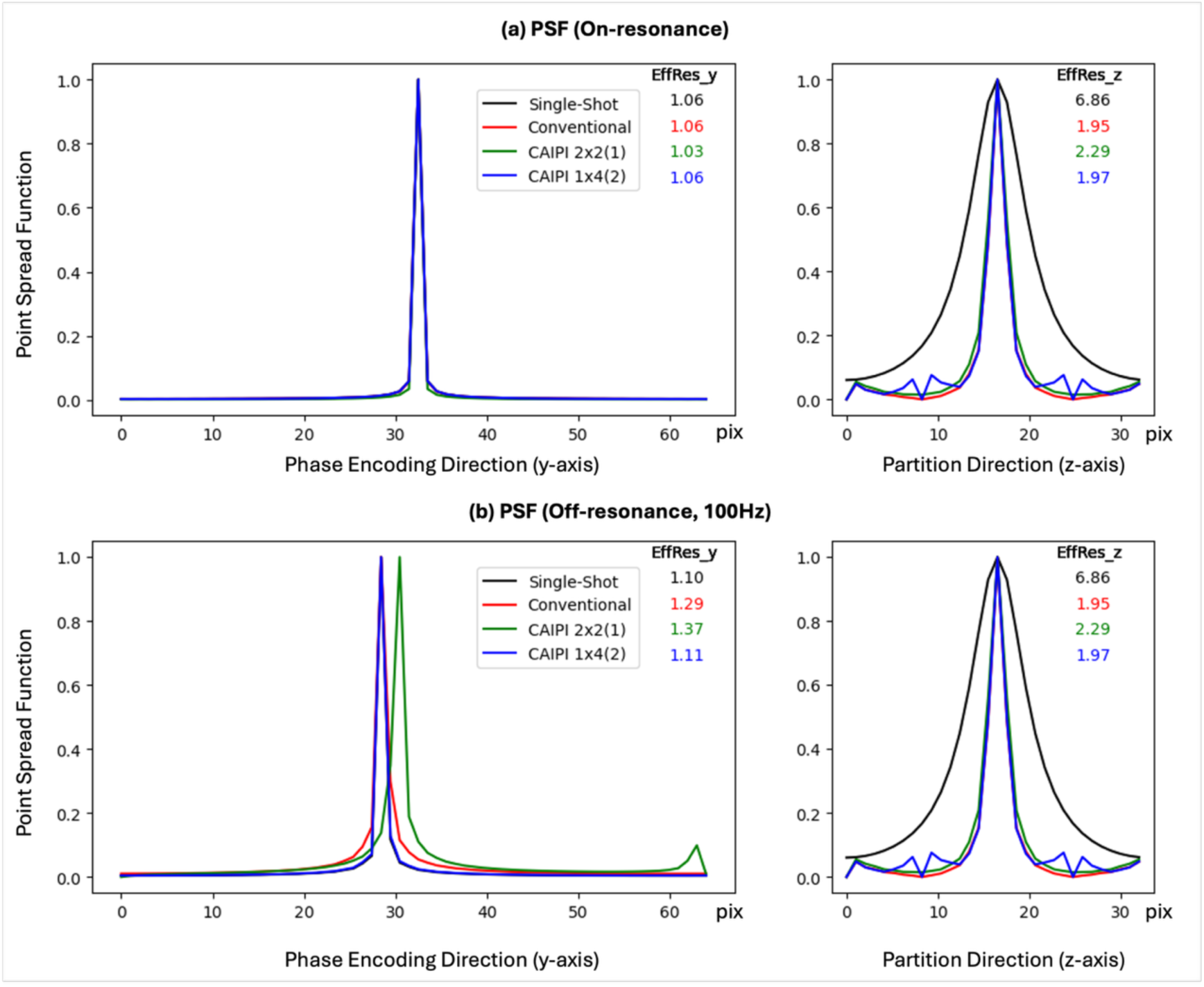
Simulated Point Spread Function (PSF) of different acquisition trajectories along the two phase-encoding directions. (a) PSF for on-resonance, and (b) PSF for off-resonance with a simulated frequency offset of 100 Hz. The single-shot PSF is included for comparison, demonstrating that segmented methods yield improved performance. The effective resolution was determined by measuring the full width at half maximum (FWHM) of the main lobe of the Point Spread Function in simulation. Effective resolution along each axis (𝐸𝑓𝑓𝑅𝑒𝑠_z_ and 𝐸𝑓𝑓𝑅𝑒𝑠_y_) is indicated in the corresponding colors for each trajectory. The simulations were performed using T2=110 ms, T2*=66 ms and a refocusing flip angle of 180° for simplicity.

In the on-resonance setting, the conventional trajectory exhibited the narrowest effective resolution in the partition direction ( 𝐸𝑓𝑓𝑅𝑒𝑠_’_ = 195 % of ideal, followed closely by 𝐶𝐴𝐼𝑃𝐼 1 × 4 (2) (197%).𝐶𝐴𝐼𝑃𝐼 2 × 2 (1) showed slightly broader resolution (229%), while the single-shot baseline exhibited substantial blurring (686%) due to its longer echo train. In the phase-encoding direction, the effective resolution was comparable across all designs (𝐸𝑓𝑓𝑅𝑒𝑠_&_ ≅ 106%).

Under 100 𝐻𝑧 off-resonance conditions, the effective resolution in the partition direction remained unchanged. In the phase-encoding direction, the single-shot scheme achieved the best resolution (𝐸𝑓𝑓𝑅𝑒𝑠_&_ = 110%) closely followed by 𝐶𝐴𝐼𝑃𝐼 1 × 4 (2) (111%). The conventional trajectory and 𝐶𝐴𝐼𝑃𝐼 2 × 2 (1) schemes exhibited greater blurring, with resolutions of 129% and 137% of ideal, respectively.

These simulations assumed a 180° refocusing pulse whereas a 120° flip angle was used in practice, which resulted in a lower signal initially but a slower signal decay over time. As a result, the actual PSFs were expected to be better than those shown here, leading to improved effective resolution in practice, although the necessary extended phase graph simulations to simulate this more accurately were beyond the scope of this work.

Both of the CAIPI variants were able to successfully reconstruct good quality navigator images in vivo, although with differing noise amplification properties. g-factor maps (**Figure 3**) for the reconstruction of the navigator images were compared between the two CAIPI variants to assess noise amplification due to undersampling within the parallel imaging reconstruction. The 𝐶𝐴𝐼𝑃𝐼 2 × 2 (1) pattern exhibited lower g-factors overall (mean = 1.15 within brain mask) with better values in central regions but elevated values near the edges, especially in inferior brain regions. The 𝐶𝐴𝐼𝑃𝐼 1 × 4 (2) trajectory, in contrast, showed higher overall noise amplification (mean = 1.32 within the brain mask) and demonstrated a wider distribution of g-factor values with less spatial homogeneity.

**Figure 3.**
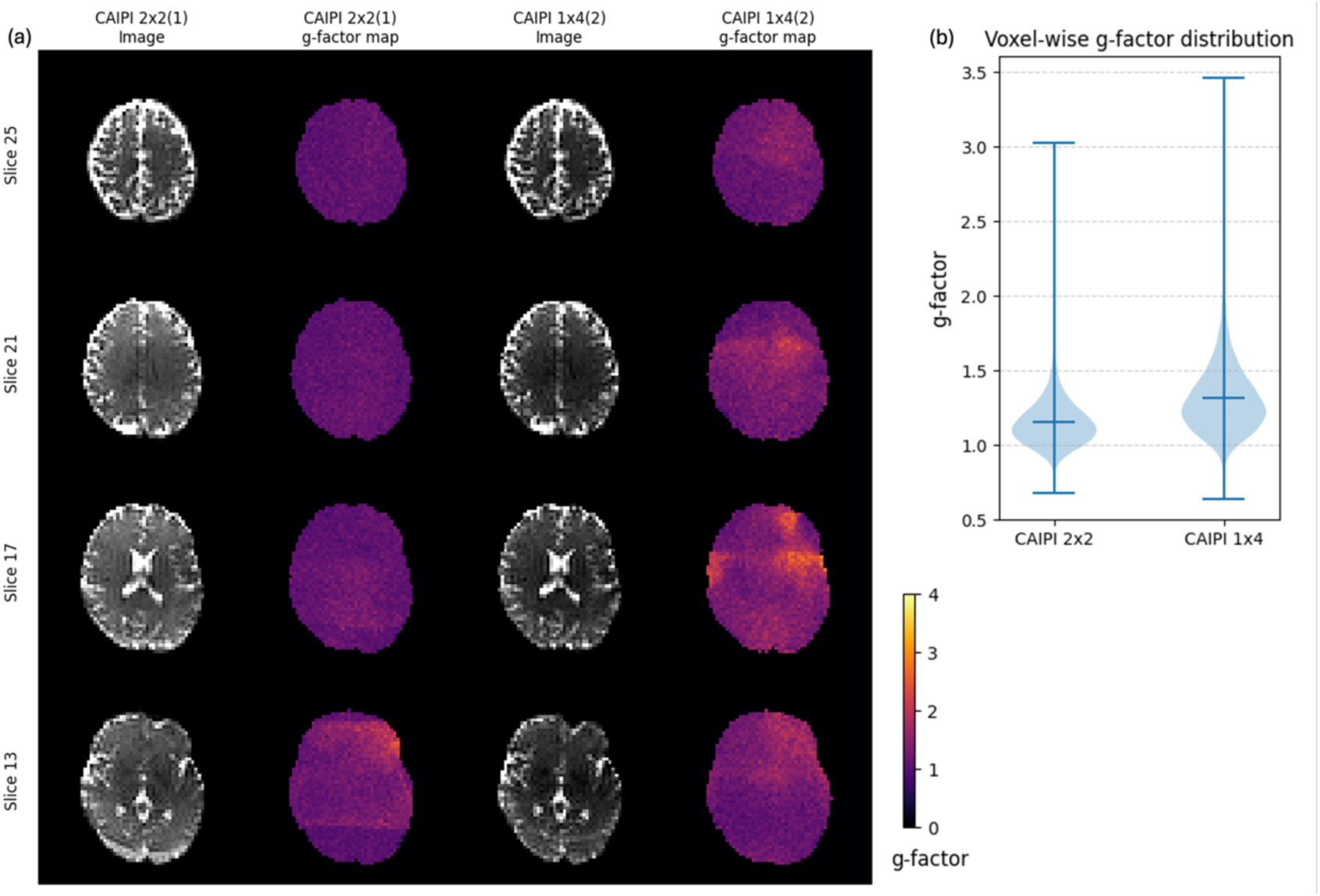
Comparison of navigator image g-factor maps and distributions for different CAIPI sampling trajectories. (a) Representative navigator axial slices (from superior to inferior) show g-factor maps for CAIPI 2×2(1) and CAIPI 1×4(2) sampling patterns. The CAIPI 2×2 (1) pattern demonstrates more uniform g-factor distributions with lower values in central brain regions (dark purple areas) while the CAIPI 1×4(1) pattern exhibits higher overall g-factors with a more heterogeneous spatial distribution (red and yellow areas). (b) Violin plot of voxel-wise g-factors from subject 2, confirming the better noise performance and greater uniformity achieved with CAIPI 2×2(1) sampling pattern.

### 3.2 Comparison of different inter-shot motion correction methods

Figure 4 shows the estimated rigid-body motion parameters, comparing results from image registration of navigator images using mcFLIRT and alignedSENSE^19^ joint estimation. The horizontal axis represents the cumulative shot index, with each complete volume comprising four shots (𝑅 = 4). The subject remained still during the first 24 shots (six volumes, three tag-control pairs), followed by cued moderate head movements. Both methods produced similar motion estimates, with only minor differences observed relative to the overall motion amplitude, indicating consistent and reliable motion estimation across frameworks.

**Figure 4:**
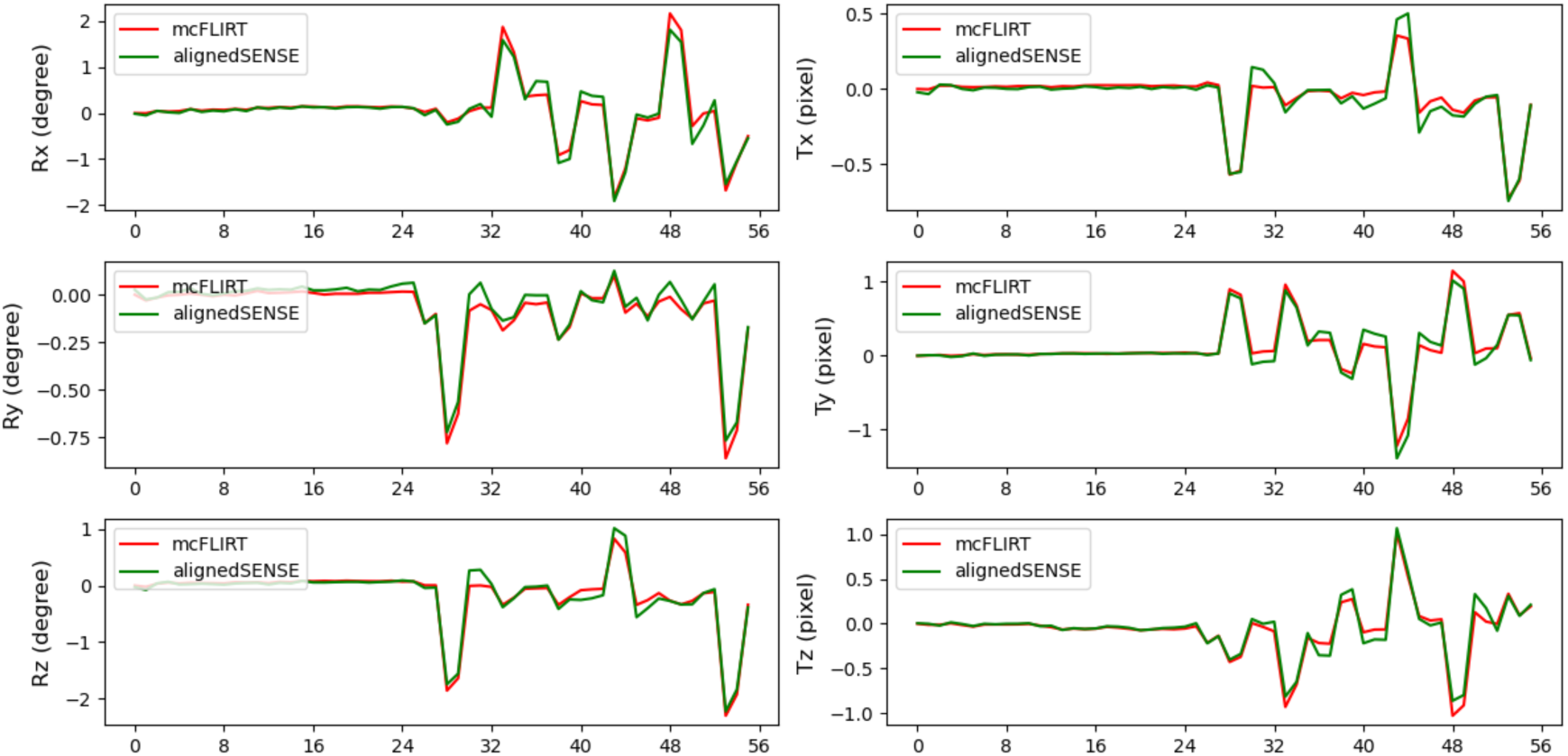
Comparison of motion parameters estimated by mcFLIRT applied to the navigator images (red) and alignedSENSE (green) for subject 4 with the CAIPI 1×4 (2) trajectory and T/C inner loop. The rotation center is set to the origin of the coordinate system.

**Table 2** presents the quantitative metrics (Corr, SSIM and tSNR) of the final corrected images, comparing the proposed inter-shot motion correction method versus alignedSENSE. The results were averaged across all five subjects, all trajectories (conventional, 𝐶𝐴𝐼𝑃𝐼 2 × 2 (1) and 𝐶𝐴𝐼𝑃𝐼 1 × 4 (2)), and both T/C interleaving strategies. The proposed inter-shot motion correction method demonstrated comparable performance to alignedSENSE across all metrics, with a modest improvement observed in the mean values of Corr (0.79 vs 0.78), SSIM (0.77 vs 0.76) and tSNR (2.02 vs 1.91). The proposed method achieved these results with substantially reduced computational time, approximately 5 times faster than alignedSENSE due to the elimination of iterative joint optimization.

**Table 2.** Correlation, SSIM, and tSNR values for different motion estimation and correction methods. The results are averaged across all five subjects, all trajectories (conventional, CAIPI 2×2 (1) and CAIPI 1×4(2)) and both tag/control interleaving strategies.

| Methods | Corr | SSIM | tSNR |
| --- | --- | --- | --- |
| AlignedSENSE | 0.7823±0.0891 | 0.7639±0.0984 | 1.9051±0.7976 |
| Self-navigation + mcFLIRT<br>(proposed method) | 0.7933±0.0962 | 0.7668±0.0911 | 2.0225±0.8724 |

Figure 5 presents the computational time breakdown for both methods. The proposed approach required only 20% of the total processing time compared to alignedSENSE. The efficiency stems from the straightforward pipeline: self-navigator reconstruction (56%), motion estimation via mcFLIRT (2%), and motion-compensated reconstruction (42%). In contrast, alignedSENSE’s iterative joint optimization dominated processing time, consuming 389% of the proposed method’s total time for motion estimation and 108% for image estimation.

**Figure 5.**
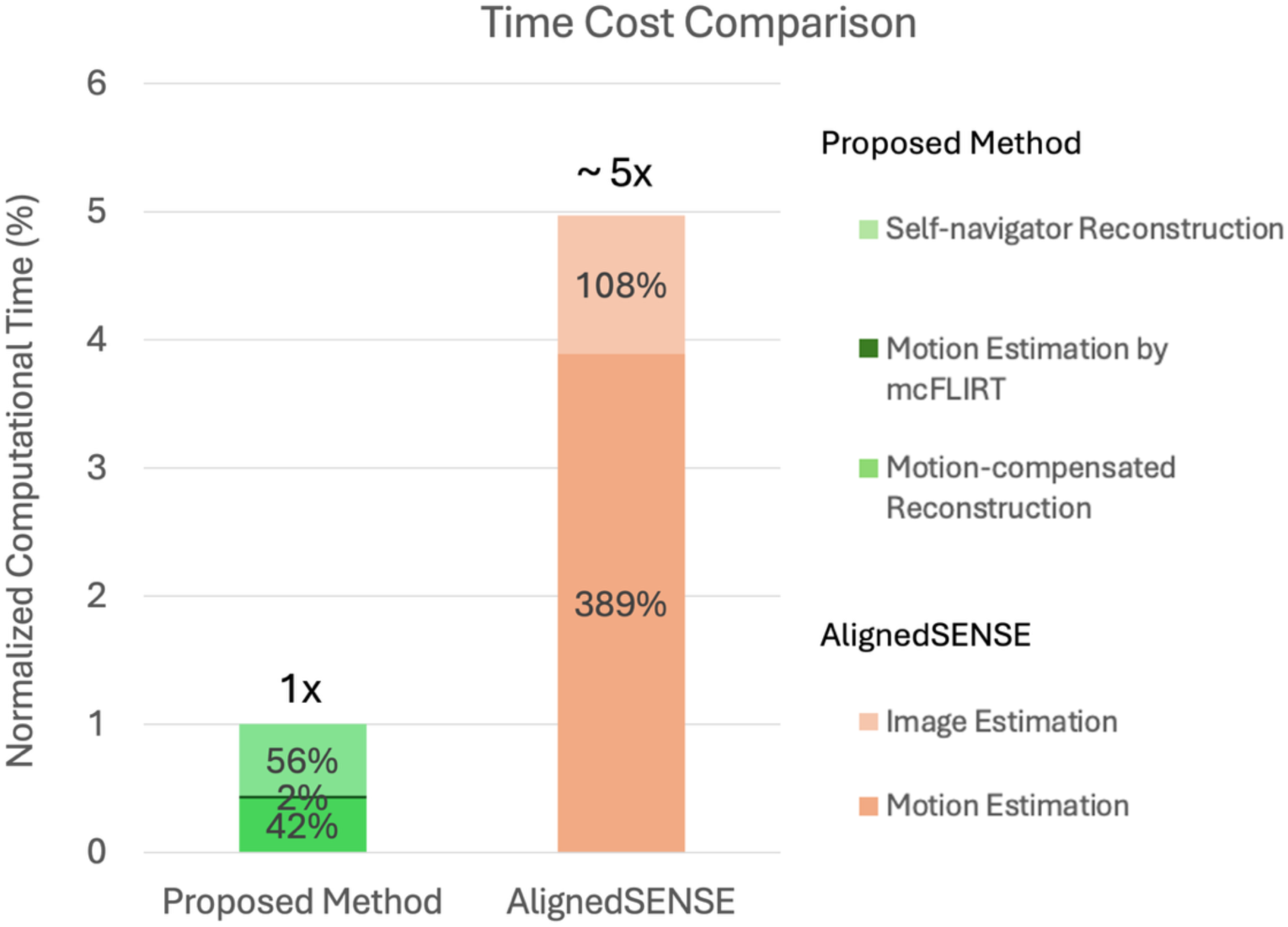
Computational time comparison between the proposed method and alignedSENSE on the same hardware. The bar chart shows the normalized computation time for different processing components, with the proposed method set as the reference (1×). The proposed method consists of three main stages: self-navigator reconstruction (56% of total time), motion estimation by mcFLIRT (2% of total time), and motion-compensated reconstruction (42% of total time). AlignedSENSE requires about 5× longer processing time, with the majority spent on iterative joint image and motion estimation (108% and 389% of proposed method’s total time respectively).

### 3.3 Comparison of different trajectories and T/C interleaving strategies

Figure 6 and Figure 7 present the temporally averaged perfusion images of Subject 4 in the axial and sagittal planes, respectively, comparing motion-free reference images, motion-corrupted images and results after inter-volume and inter-shot motion correction across different trajectories and T/C interleaving strategies. Comparing the motion-free and motion-corrupted images revealed that inter-shot motion introduced strong artefacts, particularly near gray matter-CSF boundaries. Inter-volume motion correction mitigated these artifacts somewhat, but residual artifacts remained evident. In contrast, the proposed inter-shot motion correction greatly reduced the impact of these artefacts, restoring perfusion signals close to the motion-free reference.

**Figure 6.**
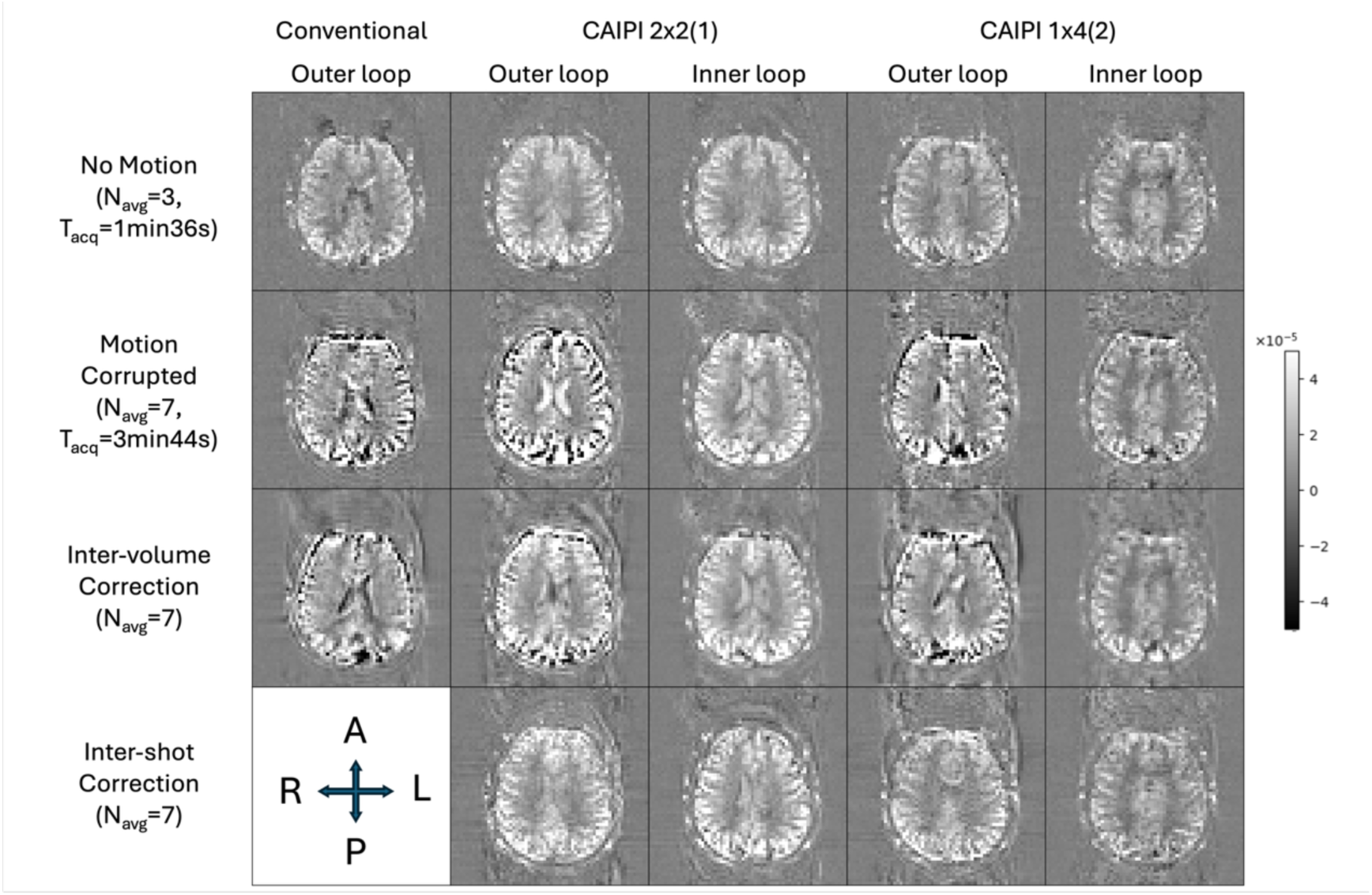
Visualization of perfusion images for subject 4 (axial view). The perfusion images are generated by subtracting the magnitude of the tag images from the control images and averaging over time. Both positive and negative signals are shown here to highlight the appearance of artifacts. “No Motion” indicates the perfusion images obtained when the subject remained still (3 tag-control pairs), while “Motion Corrupted” shows the averaged images from the whole experiment, including 4 tag-control pairs captured when the subject moved their head according to instructions. “Inter-volume Correction” refers to post-processing image registration across volumes, whereas “Inter-shot Correction” describes the proposed method that addresses inter-shot motion. Note that inter-shot motion correction cannot be performed using the conventional trajectory due to the highly asymmetric k-space sampling within each shot. The results demonstrate that the proposed inter-shot correction, especially when combined with T/C inner-loop interleaving, substantially reduces motion-related artifacts and improves perfusion image quality.

**Figure 7.**
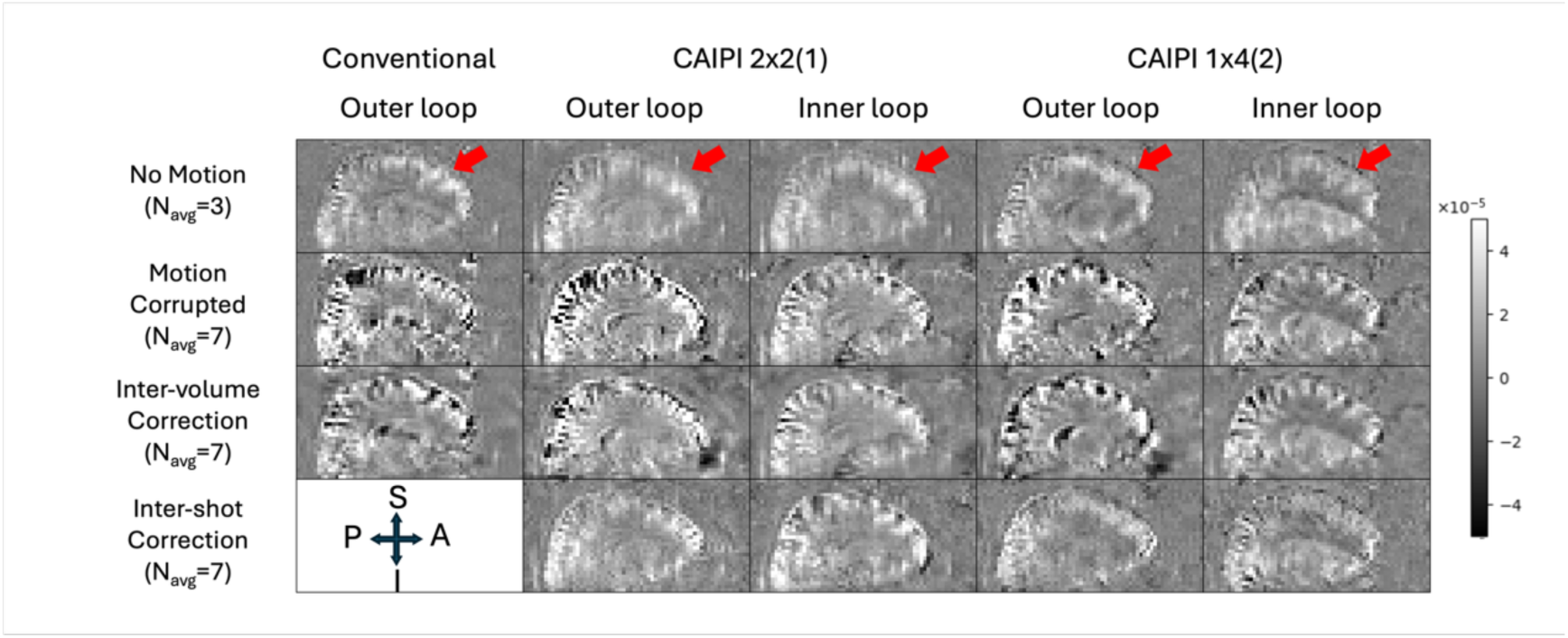
Visualization of perfusion images for subject 4 (sagittal view). Arranged as per Figure 5. Blurring along the partition (z) direction is highlighted with red arrows, most apparent for the CAIPI 2×2 (1) trajectory due to the larger number of refocusing pulses. In contrast, CAIPI 1×4 (2) preserves resolution comparable to the conventional trajectory, while remaining applicable to the proposed inter-shot motion correction.

Regarding T/C interleaving, the outer loop consistently exhibited more severe motion artifacts, especially near the brain edges, and the motion-corrected images were noisier compared to using T/C inner loop. Among the trajectories, 𝐶𝐴𝐼𝑃𝐼 2 × 2 (1) appeared to provide higher signal intensity in the axial plane in the image center, but was considerably blurrier in the sagittal view (Figure 7).

Figure 8 quantitatively compares inter-volume and inter-shot motion correction across all trajectories in five subjects using Corr, SSIM, and tSNR metrics. Results were grouped by sampling trajectory (Conventional, 𝐶𝐴𝐼𝑃𝐼 2 × 2(1), 𝐶𝐴𝐼𝑃𝐼 1 × 4(2)) and T/C interleaving strategy (outer loop vs. inner loop). Across all CAIPI trajectories with T/C inner loop, the proposed inter-shot motion correction method consistently outperformed inter-volume correction. On average, the inter-shot correction method improved correlation coefficients by 12.3%, SSIM by 4.5%, and tSNR by 40.1% compared to inter-volume correction. Although the metric differences did not always reach significance, a consistent trend favoring inter-shot correction was observed across all settings, including T/C outer loop.

**Figure 8.**
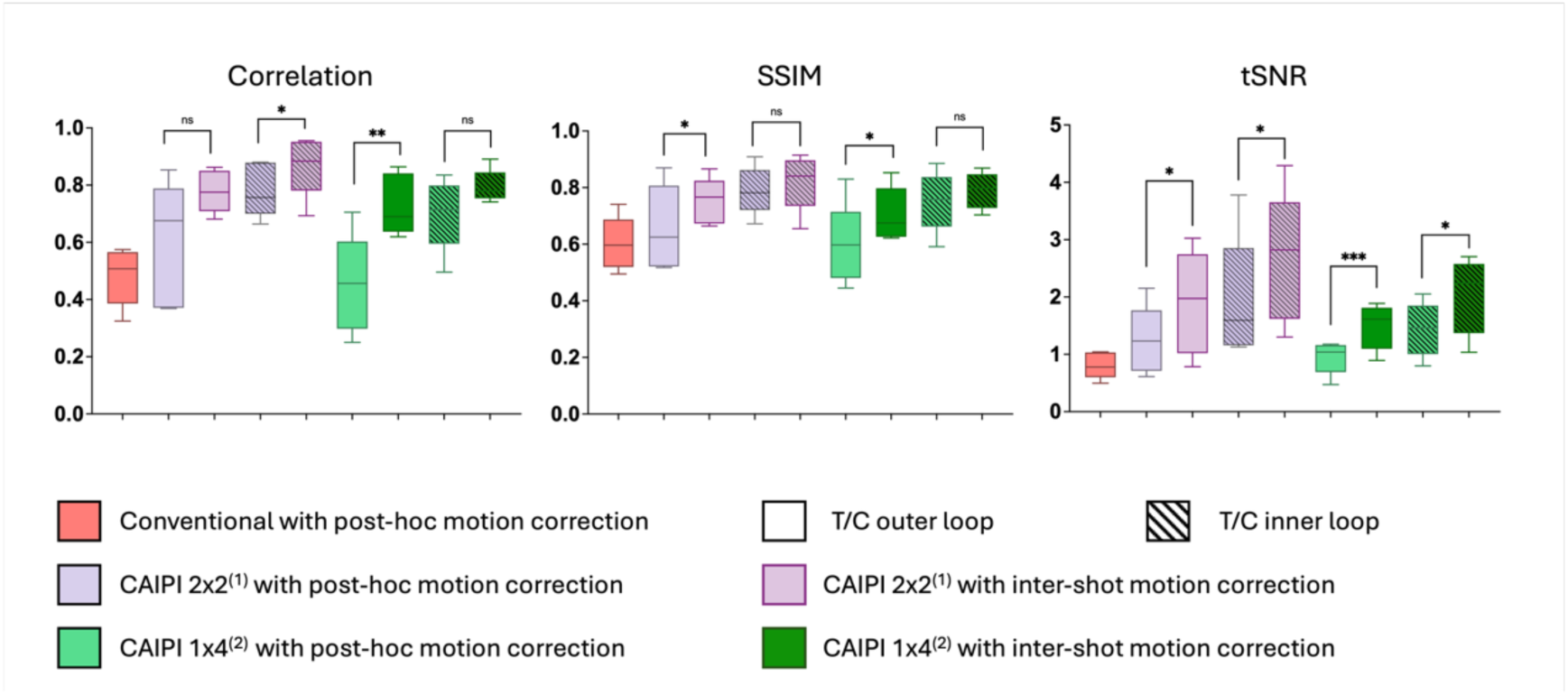
Quantitative comparison of post-hoc motion correction and the proposed inter-shot motion correction across various metrics, including correlation coefficients, SSIM, and tSNR for all trajectories. A Paired t-test was conducted to compare values before and after motion correction. Significance levels: ns (P > 0.05), * (P ≤ 0.05), ** (P ≤ 0.01), *** (P ≤ 0.001).

Comparing inner vs. outer T/C loop configurations within each trajectory showed that the T/C inner loop consistently yielded better image quality and motion correction performance. For 𝐶𝐴𝐼𝑃𝐼 2 × 2(1), the inner loop improved performance by 20.0% for correlation, 14.3% for SSIM and 46.4% for tSNR compared to the outer loop. Similarly, 𝐶𝐴𝐼𝑃𝐼 1 × 4(2) showed improvements of 26.7% for correlation, 18.4% for SSIM and 42.2% for tSNR. These results highlighted the advantage of acquiring T/C pairs in close temporal proximity to minimize interleaving-related motion effects.

The conventional trajectory, which used a T/C outer loop and did not support inter-shot correction due to half k-space coverage per shot, exhibited the lowest performance across all metrics (Corr: 0.48, SSIM: 0.60 and tSNR: 0.81). In contrast, both 𝐶𝐴𝐼𝑃𝐼 2 × 2(1) and 𝐶𝐴𝐼𝑃𝐼 1 × 4(2) trajectories achieved substantially higher scores, even with inter-volume correction only. Notably, 𝐶𝐴𝐼𝑃𝐼 2 × 2(1) with inter-shot correction and T/C inner loop delivered the best overall performance (Corr: 0.87, SSIM: 0.82, tSNR: 2.67).

To further assess the statistical significance of the observed differences, a multi-way ANOVA was conducted on the combined metrics across all CAIPI trajectories (excluding conventional trajectory), with factors including trajectory type, T/C interleaving strategy, and motion correction method. T/C interleaving strategy showed an extremely significant effect (F = 28.58, p < 0.001), providing strong statistical evidence that the inner loop configuration consistently outperforms the outer loop approach. Trajectory type also demonstrated a significant effect (F = 10.24, p = 0.003), confirming meaningful performance differences between the two CAIPI sampling variants. Finally, the choice of motion correction method showed a significant effect (F = 33.58, p < 0.001), confirming the benefits of inter-shot correction over conventional inter-volume methods.

## Discussion

In this study, we proposed a self-navigated inter-shot motion correction framework for segmented 3D-GRASE ASL imaging, enabled by CAIPI sampling trajectories. Our results demonstrated that this approach improved motion robustness and overall image quality compared to a conventional inter-volume correction method, and achieved comparable or better performance than joint estimation methods such as alignedSENSE, and with less computational resources. In addition, we evaluated the impact of tag-control interleaving strategies, providing experimental evidence supporting the use of an innermost T/C loop in segmented 3D acquisitions.

The use of CAIPI sampling played a central role in the effectiveness of our method. CAIPI sampling enabled each shot to be reconstructed individually with sufficient image quality for reliable motion estimation, whereas conventional segmented sampling patterns (such as half k-space coverage per shot) did not provide adequate data for robust per-shot reconstruction. Although the additional phase blips required in both y and z directions for CAIPI slightly compromise the PSF, CAIPI 1×4(2) still demonstrated excellent resolution under on-and off-resonance conditions while CAIPI 2×2(1) exhibited more blurring in the partition direction due to the larger number of refocusing pulses required, which can be observed in Figure 7.

The proposed motion correction pipeline leveraged the full k-space coverage of CAIPI sampling to reconstruct per-shot navigator images without the need for a separate navigator acquisition, which would add acquisition complexity, perturb the magnetization evolution and usually result in lower-resolution navigator images^30^. Our approach allowed accurate rigid-body motion estimation for each shot using conventional registration tools, e.g. mcFLIRT^24^. This was an advantage over conventional inter-volume registration performed in post-processing, which cannot correct for inter-shot motion, whilst also avoiding potential interpolation-induced blurring in post-processing by incorporating motion correction directly into the reconstruction. Our experimental results demonstrated that inter-shot motion correction consistently improved correlation, SSIM, and tSNR metrics across all tested trajectories, particularly when used in combination with an innermost T/C loop.

The approach of reconstructing individual shots for image-based navigation has been investigated in previous work for various acquisition schemes^31,32^. However, the key insight here was that the CAIPI sampling design enabled a simpler implementation than joint estimation methods like alignedSENSE. While alignedSENSE can handle cases where individual shots cannot be reliably reconstructed and provides a general framework for motion-corrupted parallel imaging, it requires iterative joint optimization with careful regularization tuning and is potentially more sensitive to initialization. In contrast, for the specific case of CAIPI sampling with moderate acceleration (R=4), the well-conditioned per-shot reconstruction quality eliminated the need for complex joint simultaneous motion estimation by aligning all navigator images to a single reference, requiring no additional post-processing stages while achieving similar image quality with substantially reduced computational time.

The order of the tag-control interleaving loop also had a significant impact on motion robustness. While consensus guidelines have recommended the use of an inner-loop configuration, our study provided the experimental validation of this recommendation in the context of segmented 3D-GRASE. We observed that inner loop T/C pairing consistently reduced motion artefacts and yielded cleaner perfusion maps, especially in cortical regions where motion sensitivity is highest.

Despite promising results, this study still has several limitations. First, all experiments were conducted on healthy volunteers with instructed moderate motion. In practice, motion patterns can vary significantly between individuals. Further validation in patient populations and under more natural, uncontrolled motion conditions is necessary. Second, the acceleration factor of each navigator used in this study was limited to R = 4, which allowed high-quality self-navigator reconstruction. If a higher degree of readout segmentation is desired, to further reduce blurring artefacts or push to higher spatial resolutions, for example, then further work is needed to assess the robustness and limits of the proposed framework under more aggressive undersampling conditions. Third, the current implementation requires offline reconstruction processing. While the computational efficiency of the proposed method makes real-time implementation more feasible than iterative approaches like alignedSENSE, integration into scanner console software would require further development to enable online motion correction.

The principles underlying this approach are generalizable beyond segmented 3D-GRASE acquisitions. The framework could potentially be extended to other multi-shot sequences such as stack-of-spirals or segmented EPI, provided each shot allows sufficient k-space coverage for reliable navigator reconstruction.

## Conclusions

We have presented a self-navigated inter-shot motion correction framework for segmented 3D-GRASE ASL enabled by CAIPI sampling, which allows robust per-shot navigator reconstruction and accurate rigid motion estimation. This approach effectively mitigates inter-shot motion artifacts, outperforming conventional inter-volume correction and matching or exceeding joint estimation methods like alignedSENSE, with lower computational complexity. Additionally, we experimentally validated the superior motion robustness of tag-control interleaving placed in the innermost loop. These findings highlight the potential of the proposed method to improve ASL image quality in the presence of motion, paving the way for more reliable clinical and research applications.

Future work will explore higher acceleration factors and patient cohorts to further establish clinical utility.

## Data Availability

We are currently unable to share in vivo data due to data protection issues, although our center is actively working on a solution to this.

## Acknowledgements

This work conducted in the Oxford Centre for Integrative Neuroimaging was supported by core funding from the Wellcome Trust (203139/Z/16/Z and 203139/A/16/Z) with additional support from the NIHR Oxford Biomedical Research Centre (NIHR203311) and the Oxford Health Biomedical Research Centre (NIHR203316). M.H. is supported by the Jardine Foundation. M.C. is supported by the Canada Research Chairs Program. P.J. is supported by the Vivensa Foundation and the NIHR Oxford Biomedical Research Centre. T.O. and J.W. were supported by a Sir Henry Dale Fellowship jointly funded by the Wellcome Trust and the Royal Society (220204/Z/20/Z). For the purpose of open access, the author has applied a CC BY public copyright license to any Author Accepted Manuscript version arising from this submission.

## Data Availability Statement

The code for the proposed motion correction pipeline can be found here: https://github.com/LoicHmh/asl_intershot_motion_correction. Data underlying the plots in the figures in this paper can be found here <LINK to be added upon publication>. We are currently unable to share in vivo data due to data protection issues, although our center is actively working on a solution to this.

## Conflict of Interest Statement

PJ is the Editor-in-Chief of Magnetic Resonance in Medicine. In line with COPE guidelines, he recused himself from all involvement in the review process of this paper, which was handled by an associate editor. He and the other authors had no access to the identities of the reviewers.

## Notes

### Competing Interest Statement

The authors have declared no competing interest.

### Author Declarations

The Clinical Trials and Research Governance Department of the University of Oxford gave ethical approval for the technical development protocol used in this work.

